# The Acceptability of Self-Collected Samples for STI Testing: A Qualitative Study Among Adults in Rakai, Uganda

**DOI:** 10.1101/2023.02.17.23286055

**Authors:** Yasmin P. Ogale, M. Kathryn Grabowski, Proscovia Nabakka, William Ddaaki, Rosette Nakubulwa, Neema Nakyanjo, Fred Nalugoda, Joseph Kagaayi, Godfrey Kigozi, Julie A. Denison, Charlotte Gaydos, Caitlin E. Kennedy

## Abstract

**Introduction:** Self-collected samples (SCS) for sexually transmitted infection (STI) testing have been shown to be feasible and acceptable in high-resource settings. However, few studies have assessed the acceptability of SCS for STI testing in a general population in low-resource settings. This study explored the acceptability of SCS among adults in south-central Uganda.

**Methods:** Nested within the Rakai Community Cohort Study, we conducted semi-structured interviews with 36 symptomatic and asymptomatic adults who self-collected samples for STI testing. We analyzed the data using an adapted version of the Framework Method.

**Results:** Overall, participants did not find SCS physically uncomfortable. Reported acceptability did not meaningfully differ by gender or symptom status. Perceived advantages to SCS included increased privacy and confidentiality, gentleness, and efficiency. Disadvantages included the lack of provider involvement, fear of self-harm and the perception that SCS was unhygienic.Most participants preferred provider-collected samples to SCS. Nevertheless, almost all said they would recommend SCS and would do it again in the future.

**Conclusion:** Despite a preference for provider-collection, SCS are acceptable among adults in this setting and support expanded access to STI diagnostic services.

**Key Questions:** *What is already known on this topic:* Timely diagnosis is critical for STI control, with testing being the gold standard for diagnosis. Self-collected samples (SCS) for STI testing offer an opportunity to expand STI testing services and are well accepted in high-resource settings. However, patient acceptability of self-collected samples in low-resource settings is not well described.

*What this study adds:* We found that SCS was acceptable to both male and female participants in our population, regardless of whether they reported STI symptoms. Perceived advantages to SCS included: increased privacy and confidentiality, gentleness, and efficiency; disadvantages included lack of provider involvement, fear of self-harm and the perception that SCS was unhygienic. Overall, most participants preferred provider collection over SCS.

*How this study might affect research, practice or policy:* Patient education addressing perceived disadvantages may increase SCS acceptability and support the use of SCS as a means to identify cases and control STIs in low-resource settings.

## Introduction

Countries in sub-Saharan Africa (SSA) generally follow a syndromic approach to manage non-HIV sexually transmitted infections (STIs). While practical and cost-effective, syndromic management is limited as diagnostic tool. ^1-4^ Self-collected samples (SCS) may be one way to address these limitations. SCS for STI testing (SCS/STI testing) occurs when individuals obtain a swab or fluid sample themselves, either within or outside the clinic, and send the specimen to a laboratory for testing. ^5^ Research in high-resource settings shows that SCS are as accurate as provider-based tests, ^6^ and that SCS/STI testing interventions are feasible and acceptable in diverse populations. ^7-16^ While not a replacement for clinic-based examination, SCS may be part of innovative efforts to expand STI case management beyond the provider-dependent, syndromic approach. ^17-20^ By allowing the patient to collect a sample themselves, studies suggest that SCS can circumvent some barriers to clinic- and/or provider-based STI case management, like stigma, access and privacy concerns. ^1, 21, 22^ For these reasons, the WHO recommended SCS as an additional approach to deliver STI testing services. ^23^

Despite their potential, SCS/STI testing interventions are rare in SSA due to a lack of cheap and accurate diagnostic tests^24^ and laboratory facilities. However, intensive scale-up of HIV care and treatment over the past 15 years has resulted in strengthened infrastructure in the region. As such, the potential for using cheaper technologies to facilitate STI etiologic testing is now becoming a viable reality, ^18^ especially in countries like Uganda. To develop effective interventions, context-specific data are required, yet few studies have considered the acceptability of SCS in a general population in low-resource settings. This qualitative study aimed to provide data on user acceptability of self-collected genital swabs for STI testing in both women and men in a low-resource setting.

## Methods

Implemented by the Rakai Health Sciences Program (RHSP), the Rakai Community Cohort Study (RCCS) is an ongoing, open community-based cohort of residents aged 15–49 years in agrarian communities, semi-urban trading centers and Lake Victoria fishing communities in south-central Uganda. ^25^ The RCCS includes the administration of a demographic and health questionnaire, as well as HIV testing for all consenting participants. Nested within the RCCS, the STI Prevalence Study (STIPS) aimed to estimate STI prevalence among 1,825 sexually active HIV+ and HIV-men and women aged 18–49 years in two communities (one inland and one fishing), from May to October 2019. ^26^ STIPS participants were tested for Trichomonas vaginalis (TV) (in the field), syphilis (screening in the field; samples tested in the lab), N. gonorrhoeae (NG), C. trachomatis (CT), and herpes simplex virus type 2 (HSV-2) (samples tested in the lab). To this end, three provider-collected penile-meatal swabs were obtained for all male STIPS participants who consented to STI testing. Because we were interested in men’ s experience with SCS, a fourth, self-collected swab was obtained from a sub-sample of men (n=40); it is from this sub-sample that we recruited the male study participants for our qualitative interviews (n=15).

Three self-collected vaginal swabs were obtained for all female STIPS participants who consented to STI testing (provider-collected samples were not obtained for females); it is from this sample that we recruited the female study participants for our qualitative interviews (n=21). All participants who self-collected samples received instructions from a same-gender provider before sample collection and were then given privacy to self-collect. Interviews were conducted after participants received their HIV, TV and syphilis screening results but before their NG, CT and HSV-2 results. Individuals who tested positive for STIs were provided treatment by RHSP according to the Ugandan National Clinical Treatment Guidelines for Sexually Transmitted Infections.

This qualitative study was conducted among 36 adults—15 men and 21 women—from the STIPS rural, inland community who self-collected a sample in STIPS. We selected participants based on gender and self-reported symptom status, with 9/15 (60%) men and 15/21 (71%) women reporting at least one STI-related symptom in the last six months. Symptoms included: genital ulcer, genital discharge, frequent urination, painful urination, pain during intercourse, bleeding during intercourse, lower abdominal pain, genital warts, and for females: thick and/or colored vaginal discharge, vaginal itching and unpleasant vaginal odor. We conducted semi-structured interviews that explored participants’ experiences and preferences related to SCS/STI testing. Interviews were conducted in a private location of the participant’ s choosing. RHSP social and behavioral scientists conducted all interviews in Luganda. The interviewers and study lead debriefed after each interview.

Interviewers transcribed and translated interviews into English. We then imported the data into MAXQDA 2018^27^ for review and initial analysis. Our analysis methods were adapted from the Framework Method. ^28^ First, we reviewed the interviews in MAXQDA to familiarize ourselves with the data. Second, we developed an analytic framework comprised of categories that were informed by our interview guide and research questions. We used this framework to index the interviews. Third, after all interviews were indexed, we charted the data into a framework matrix in Excel. Fourth, we conducted open-ended coding, followed by focused coding, ^29^ to identify prominent themes within each category. Prominent themes were defined by the depth of discussion any one participant provided on the topic, prevalence across participants and ‘keyness’ in relation to our research questions. ^30^ Fifth, we compared the themes by gender and symptom status to assess for any meaningful differences. Finally, we discussed our findings among the research team, including interviewers and co-investigators, to ensure clarity and cohesion.

### Ethical Consideration

We obtained ethical clearance from the Johns Hopkins Medicine Institutional Review Board (IRB00204691; July 9, 2019), the Uganda Virus Research Institute Research and Ethics Committee (GC/127/19/07/709; July 19, 2019) and the Uganda National Council for Science and Technology (HS364ES; June 6, 2019). Interviewers obtained written consent from participants prior to data collection.

## Results

Below, we present participants’ experiences and preferences related to SCS/STI testing. Participants are described by their gender (M: male; F: female) and symptom status (S+: self-reported symptoms; S-: no self-reported symptoms). Table 1 provides illustrative quotations identified by letters to match the corresponding themes in the text below.

**Table 1.** Illustrative quotations by theme. Participants are described by their gender (M: male; F: female) and symptom status (S+: self-reported symptoms; S-: no self-reported symptoms)

| Section header and corresponding text reference | Quotation (Participant description) |
| --- | --- |
| Overall experience |  |
| A | *To be honest, me, I didn't find any problem with it...It was easy to me and I was very happy about it. (M, S-)* |
| B | *This time, we were given a chance to do it by ourselves without any difficulty.... I felt so good I was not scared at all; I did everything as instructed by the musawo <doctor> and I was able to collect the sample myself. (F, S+)* |
| C | *[I] am satisfied [with] being checked by a musawo, I don't like self-testing. (F, S+)* |
| SCS advantages |  |
| D | *I prefer doing it [collecting a sample] myself...If a musawo collected from me and touches my penis, somehow, I will feel shy. (M, S+);* |
| E | *The good thing is that if I self-collect there is nothing like obuswavu <showing your nakedness> compared to when the health worker collects it....when the musawo is collecting the sample I must squat and then the musawo will see my private parts when removing the swab... I prefer collecting it myself. (F, S-)* |
| F | *Personally the issue I have noticed there with the musawo collecting the sample is...[because] you were not given proper notice, so probably you came when you have not groomed or prepared yourself well. That is the problem I see.... [laughs] the musawo may find when you are somehow dirty [laughs].... You might come when you have not cleaned up and she says, get ready am collecting the sample and you get embarrassed because you came not well prepared. (F, S+)* |
| G | *[Self-collecting] is good, and [secrets don't] spread because it is you that takes it off and give it to the health worker and it stops at you two....[Self-collection] will also continue to keep secrets because it remains with just you. (M, S-)* |
| H | *For those people who are afraid of approaching a health worker to tell him/her the truth; it will be good because they will be self-testing and doing everything by themselves...everyone has their own secrets that they are hiding. (F, S+)* |
| I | *You may find someone [a provider] who presses [the swab] so hard...but if it is you...[and] you get it yourself very well and find that you do not feel the pain like [when] the musawo does [it]. (M, S-)* |
| J | *Musawo, I know my body; the musawo may insert it far. [laughs]...I would be thinking that what if she pierces me. (F, S+)* |
| K | *[Because] you can test yourself, it helps you to save money, time say that you would have used from here for example to Kalisizo [the neighboring hospital]. (M, S+)* |
| SCS disadvantages |  |
| L | *The health worker is more experienced in carrying out these tests and gets to know the results very fast. As for me, I will be there debating whether I carried out the test in the right way. (M, S-)* |
| M | *I might [use the swab] wrongly or insert it wrongly, I might not know exactly how best to insert it and how far it should go, however a musawo knows how best to insert and how far it should go and how best to remove it. (M, S+);* |
| N | *My worry is that [I] may insert it wrongly and hurt the uterus which may not be the case when the musawo does it because she knows how everything is. (F, S+);* |
| O | *Personally, I would prefer the health worker to collect...she is a health worker, she can't insert it as if she is going to kill you. (F, S+)* |
| P | *What causes me to fear is [that the provider] puts on gloves, [inaudible] and yet he has told me to do it with bare hands. Which means I can come when I have cleaned up myself, but someone else may come from the garden, has been digging and then starts from there. Now don't you see his hands, if they have germs and then he touches his genital areas...And it is not good for them. (M, S-)* |
| Preference and future use |  |
| Q | *I would use [SCS in the future] because sexually transmitted infections don't just come in a particular time and stop – they come any time – so I would like to keep using this method so that I can know where I stand. (M, S-)* |

### Overall experience

In terms of their experience self-collecting a sample, almost all participants reported ‘never [feeling] bad’ during the collection process and that they had ‘no problems’ with it [A].

Participants said the SCS instructions given by the provider before self-collection were helpful [B]. Overall, participants found the SCS process to be comfortable physically. The majority stated that they ‘never felt any pain’ during sample collection. A few men did indicate a minor discomfort when taking the swab but described it as ‘some little pain’ that was ultimately ‘bearable’ (M, S+). While participant responses were generally positive, two symptomatic women did not appreciate the SCS experience: the first woman did not feel comfortable because it was a new method, while the second simply did not like it [C].

### SCS advantages

Advantages of SCS included privacy and confidentiality and gentleness. SCS was also more efficient, as it addressed challenges due to transportation, time and money, which make clinic attendance difficult.

Regarding privacy and confidentiality, some participants felt that SCS removed feelings of shyness and embarrassment associated with undressing in front of a provider. This sentiment was expressed by both men [D] and women [E].

Some participants also liked SCS because it avoided embarrassment caused by being ‘dirty,’ which related to being ungroomed, unkempt or unclean in the genital area [F].

A few men wanted to avoid this embarrassment as a professional courtesy to the provider. As one man described, a patient may come when they are ‘*munda oyo tasawayo* <not shaven>;’ this could ‘for sure…scare the health care worker.’ He later stated that self-collecting a sample was best as it would avoid disturbing the provider in such a way (M, S+).

Confidentiality was another perceived advantage to SCS. Some participants, including both men and women, felt that SCS was more confidential than provider-collection. Participants described a local environment of mistrust and rumor mongering: ‘people in the community are rumor mongers,’ explained one woman, ‘they tell everyone.’ (F, S-) Participants therefore valued confidentiality and the ‘keeping of secrets;’ participants felt that SCS allowed them to maintain the secret that they participated in STI testing [G]. SCS was also advantageous for those who were afraid to discuss their private matters with a provider [H].

Some participants felt that SCS was gentler than provider-collection. This was especially true among participants who reported symptoms. Both men and women feared that the provider would inflict pain when taking a sample [I]. Some participants felt that SCS would be less painful than provider-collection because the patient ‘knows their own body,’ unlike the provider [J].

Finally, some participants felt that SCS was more efficient, especially if used at home, and would save time and money [K]. Some participants also perceived SCS to be faster than provider-collection, where clinic waiting times could cause delays.

### SCS disadvantages

Reported disadvantages of SCS included the lack of provider involvement (and thereby, their training and expertise) in the collection process, fear of self-harm and the perception that SCS was unhygienic.

Regarding provider involvement, some participants worried that they may collect a sample poorly if they were to collect it in the absence of a provider. Participants felt that the provider was trained and more experienced, and as such, would carry out the process better than they would themselves [L, M].

A perceived risk of self-harm was another disadvantage of SCS. Some participants were afraid of hurting themselves if they took the sample [N, O].

Finally, some participants were concerned that SCS was unhygienic: because they do not wear gloves, participants were afraid of spreading germs in their genital area during self-collection; provider-collection was more sanitary because providers wore personal protective equipment [P].

### Preference and future use

When asked for their ultimate preference, more participants preferred provider-collection over SCS (18/36 [50%] for provider versus 13/36 [36%] for SCS; 5/36 [14%] with no preference). This was true regardless of symptom status or gender. Of those that preferred SCS (n=36), however, women—especially those reporting symptoms—were slightly more likely to prefer SCS than men (9/21 [43%] of women selected SCS versus 4/15 [27%] of men).

Nevertheless, we found that almost all participants would recommend SCS to others, whether it be their friends, family or peers. We also found that almost all participants, except one woman, were willing to use SCS again in the future. Many recognized the utility of SCS as a means to receive an STI diagnosis and valued the opportunity to ascertain their disease status again in the future [Q].

While almost all participants were willing to use SCS again, one woman said she would not use SCS again out of fear that community members would spread rumors about her because she tested for STIs.

## Discussion

In this qualitative study in south-central Uganda, we found that SCS was acceptable to both male and female participants, regardless of whether they reported STI symptoms. Overall, participants reported a positive experience with self-collection. Advantages of SCS included confidentiality, privacy, comfort and efficiency. Disadvantages included a lack of provider involvement, fear of self-harm, and the perception that SCS was unhygienic. Most participants said they preferred provider-collection for STI testing. However, with one exception, all participants stated that they would recommend SCS to others and would use SCS again in the future.

Data on the acceptability of self-collected genital swabs for STI testing in a general population in low-resource settings, particularly in SSA, are rare. Our findings corroborate previous studies in Rakai, which demonstrated that self-administered vaginal swabs were valid and acceptable methods to screen for STIs among women, and urine samples were acceptable to both women and men. ^31-34^ Our findings also agree with those of a systematic review by Paudyal et al. on patient experiences obtaining self-samples to diagnose STIs. ^13^ While the review covered a variety of self-collection methods (not just genital swabs) and included only two studies from low-resource settings, it evaluated the same STIs as our study, and found that the majority of adults accepted SCS and found it to be an ‘easy’ procedure. The review also found that privacy and safety were the most common concerns adults had about SCS.

While more specific to women, we can draw comparisons between our data and data on the acceptability of self-collected swabs for HPV testing in SSA. Two studies from Uganda have examined acceptability of self-collection of HPV samples. A quantitative study of women in a low-resource community in Kampala found that more than 80% of participants were willing to collect their own HPV samples. ^35^ However, in that study, SCS was delivered by a provider to the participant’ s home (and SCS was conducted there, too); therefore the high observed acceptability could have been due to the location of sample collection (i.e. at home), the mode of delivery (i.e. by a provider), the collection method (i.e. SCS), or some combination of the three. Despite this, the study did identify some barriers to self-collection, including embarrassment due to a lack of privacy (in the home/community), worry of collecting incorrectly and older age.

Likewise, a mixed methods study conducted in India, Nicaragua and Uganda found that 75% of all women felt SCS was easy, though initial concerns included hurting themselves (52%) and getting a bad sample (24%).^36^ Women also reported an unwillingness to touch the genital region; similar to our study, participants also valued sanitation, privacy and cleanliness. ^36^

To our knowledge, this is the first study to explore the acceptability of self-collected penile-meatal swabs among men in a low-resource setting. We were surprised that SCS acceptance among men in our population was not higher. We expected that most men would rather self-collect to avoid undressing and exposing themselves in front of a provider. We also expected that the majority of men would prefer SCS, citing its flexibility to test during non-clinic hours, as has been observed in studies on the acceptability of HIV self-testing among men. ^37, 38^ We recommend researchers continue to explore SCS acceptability among men in diverse settings.

Finally, we were initially surprised by our finding that both men and women preferred a provider-collected sample over SCS, despite indicating that SCS was acceptable. The aforementioned review found that SCS was preferred to provider-collection. ^13^ A study assessing the acceptability of self-collected penile swabs among men in the U.S. also found that 77% of participants preferred a self-collection over attending a clinic. ^39^ Data on HPV self-sampling also found that while acceptability was high, participants’ preference for SCS versus provider-collection was mixed: in a systematic review among women in mostly high-resource settings, about half of the included studies showed that participants preferred SCS, while the other half showed that women preferred provider-collection because they lacked confidence in their ability to self-collect a sample correctly. ^40^ In ten of the 23 included studies, women felt that provider sampling was more reliable than SCS. Data from other settings in SSA, too, show that our findings are not unexpected: women in SSA reportedly preferred provider sampling to HPV self-sampling, or preferred having a provide present during the process, because they feared hurting themselves when self-collecting^36, 41, 42^ and/or not getting a good sample. ^36, 43, 44^

This study was novel in that it explored the acceptability of SCS among a general population of women and men in a low-resource setting and provided participants the opportunity to self-collect. Because they were able to use the swabs themselves, we were able to gather detailed and practical feedback on the ease of use and their experience. Another strength of our study included the qualitative nature of data collection. By using semi-structured interviews, we were able to gather rich descriptions and a breadth of responses. Nevertheless, social desirability bias may have affected our results: because our interviewers were RHSP staff members, participants may have responded more favorably to SCS than they would have otherwise. Additionally, they may have reported a preference for provider-collection out of respect for the RCCS providers, even though confidentiality of responses was assured and reviewed during the informed consent process. We doubt these possibilities strongly influenced our results, given the fact that participants provided both advantages and disadvantages for both collection methods.

Lastly, another strength of our study was the purposeful selection of adult participants based on both gender and symptom status. This allowed us to assess if acceptability varied between users across these groups, which could help guide the development of future SCS/STI testing interventions. However, this qualitative study focused only on men and women in the inland community and we did not specifically recruit any high-risk groups, such as truck drivers, sex workers or fisherfolk. Given their mobility and sexual risk behaviors, such groups may be priority populations for SCS/STI testing services. Understanding their acceptance of SCS is critical for future program development. Age^35^ and knowledge of how to self-collect^45, 46^ have also been shown to affect SCS acceptability. Level of education and/or socioeconomic status may also influence participant preferences. ^45, 47, 48^ We did not sample based on these criteria, but recommend future studies use mixed methods to explore how such contexts could influence SCS acceptability.

In conclusion, our study found that SCS were acceptable, but concerns over taking a sample without a provider, self-harm and poor hygiene led the majority of men and women in our population to prefer provider-collection. Nevertheless, users said they would still use SCS in the future. Together, these findings suggest that SCS are an acceptable, additional approach to current STI diagnostic methods. Future health communication messages to promote SCS/STI testing can address user concerns and emphasize the perceived advantages. SSA needs a diversity of strategies to address the burden of STIs; SCS/STI testing may be one useful tool in the toolbox.

## Data Availability

All data produced in the present study are available upon reasonable request to the authors.

## Acknowledgments

We are grateful to the community members and leaders of Rakai who participated in this research. We also give special appreciation to all the RHSP staff members and in particular the Social and Behavioral Sciences team for supporting this research. Thank you to Herman Mukiibi and Frank Lukabwe for their work with data collection. We would also like to acknowledge the support of Drs. Ronald Gray and Maria Wawer.

